# Characteristics, Management And Outcomes Of Critically Ill Covid-19 Patients Admitted To Icu In Hospitals In Bangladesh: A Retrospective Study

**DOI:** 10.1101/2020.09.24.20201285

**Authors:** Ayan Saha, Mohammad Moinul Ahsan, Tarek-Ul-Quader, Mohammad Umer Sharif Shohan, Sabekun Naher, Preya Dutta, Al-Shahriar Akash, H M Hamidullah Mehedi, A S M Arman Ullah Chowdhury, Hasanul Karim, Tazrina Rahman, Ayesha Parvin

## Abstract

**Objectives:** This study aimed to analyse the epidemiological and clinical characteristics of critical COVID-19 cases and investigate risk factors including comorbidities and age in relation with the clinical aftermath of COVID-19 in critical cases in Bangladesh.

**Methods:** In this retrospective study, epidemiological and clinical characteristics, complications, laboratory results, and clinical management of the patients were studied from data obtained from 168 individuals diagnosed with an advanced prognosis of COVID-19 admitted in two hospitals in Bangladesh.

**Results:** Individuals in the study sample contracted COVID-19 through community transmission. 56.5% (n = 95) cases died in intensive care units (ICU) during the study period. The median age was 56 years and 79.2% (n=134) were male. Typical clinical manifestation included Acute respiratory distress syndrome (ARDS) related complications (79.2%), fever (54.2%) and cough (25.6%) while diabetes mellitus (52.4%), hypertension (41.1%) and heart diseases (16.7%) were the conventional comorbidities. Clinical outcomes were detrimental due to comorbidities rather than age and comorbid individuals over 50 were at more risk. In the sample, oxygen saturation was low (< 95% SpO2) in 135 patients (80.4%) and 158 (93.4%) patients received supplemental oxygen. Identical biochemical parameters were found in both deceased and surviving cases. Administration of antiviral drug Remdesivir and the glucocorticoid, Dexamethasone increased the proportion of surviving patients slightly.

**Conclusions:** Susceptibility to developing critical illness due to COVID-19 was found more in comorbid males. These atypical patients require more clinical attention from the prospect of controlling mortality rate in Bangladesh.

## 1 INTRODUCTION

The Coronavirus Disease 2019, came into limelight in early December 2019, when some cases of pneumonia were reported in Wuhan, Hubei, China, whose causes were unfamiliar. Following laboratory assessment, it was announced that these innominate cases of pneumonia were caused by a novel strain of virus belonging to the Coronavirus family and was labelled SARS-CoV-2 (Severe acute respiratory syndrome coronavirus 2) [1]. The spread of the infection is a rising, rapidly advancing circumstance and due to this whirlwind rate of spread, COVID-19 has been pronounced as a global pandemic by the WHO since March 11, 2020. As of 23rd September, more than 31.7 million positive cases of COVID-19 have been reported in 217 countries and territories with more than 975,315 deaths. COVID-19 targets the respiratory tract of humans and has similar clinical symptoms to SARS-CoV and MERS-CoV [2-4]. Typical symptoms experienced by COVID-19 positive individuals include fever, dry cough, fatigue, headache, vomiting, diarrhoea, shortness of breath, myalgia, acute respiratory distress syndrome (ARDS) related symptoms and shock [5-8]. Previous studies reported that the patients who need intensive care tend to be older in age and male, and about 40% have comorbid conditions, including diabetes, cardiac diseases, hypertension, asthma and other chronic illnesses such as liver or kidney disease [9, 10]. According to the World Health Organization, about 5% COVID-19 patients, who are severe or critically ill require admission to an intensive care unit (ICU) [11]. However, shortages of standard healthcare resources, especially ICU supports are causing the high mortality rate of critically ill patients.

The COVID-19 pandemic has imposed an enormous burden and massive challenges to the health care system, especially ICUs, across developed, developing and underdeveloped countries. Likewise, Bangladesh also falls in the category of unfortified countries due to its high population and poor health care system [12]. A total of 7% of the county’s population are senior citizens [13]. Most of these senior citizens, as well as middle-aged people in the county, have comorbidities, such as diabetes (9.7%), asthma (5.2%), hypertension (20%), cardiac disease (4.5%) and chronic pulmonary disease (11.9%), and around 1.3 to 1.5 million cancer patients in the country are vulnerable to COVID-19 [14-17]. All of these people, who belong to a vulnerable group, may require immediate hospitalisation and intensive care if they contract COVID-19 [10]. Compared to the eight worst affected countries, Bangladesh has the lowest number of COVID-19 ICU beds per 10,000 inhabitants (Supplementary Figure 1). How the health management system with its poor and limited resources is responding to and tackling critical COVID-19 patients is a matter of inordinate concern. Therefore, it is important for health and government authorities to have information on the clinical features and outcomes of COVID-19 in critically ill cases for them to address the necessities of ICU facilities’ and prepare for a possible second wave of COVID-19 in Bangladesh. Therefore, this study aims to investigate the epidemiological and clinical features, disease severity, treatment and clinical outcomes of critical COVID-19 cases in Bangladesh with the goal of portraying a bigger picture of severe clinical manifestations of COVID-19 so that the malleability Bangladesh’s health care system can be modified in terms of tackling COVID-19.

## 2 METHODS

### 2.1 PATIENTS AND DATA COLLECTION

This sample of this study comprises 168 COVID-19 patients with definite outcomes who were admitted to Chittagong General Hospital and Chittagong Medical College Hospital (COVID-19 unit) between 1^st^ April 2020 and 7^th^ August 2020. The Chittagong General Hospital and Chittagong Medical College Hospital (COVID-19 unit) are specialised hospitals that have been authorised for managing most of the critical COVID-19 patients in the country’s economic hub, namely Chattogram city. The epidemiological and demographic data for this study were obtained from the inpatients’ files. This study was approved by the IRB of the Chattogram General Hospital Ethics Committee and the patients gave oral consent regarding data collection and usage.

### 2.2 THE CRITERIA FOR ICU ADMISSION

Based on clinical symptoms, patients were divided into mild, moderate, severe and critical groups. Most of the severe or critical patients and few moderate ill patients were admitted to the ICU. Those in the severe group have respiratory distress, i.e. a respiratory rate of ≥ 30 beats per minute in a resting state and an oxygen saturation of ≤ 92% SpO2, and those in the critical group experience respiratory failure, Sepsis and shock, thus requiring mechanical ventilation, as well as the combined failure of other organs, which require ICU monitoring and treatment. The coordinative physicians were accountable for collecting this data from the patients. ARDS was defined according to the Berlin definition [18], and shock was defined according to the Sepsis-3 criteria [19].

### 2.3 REAL-TIME RT-PCR ASSAY FOR COVID-19

Whether the cases of the sample were positive with COVID-19 was confirmed via a real-time reverse transcription polymerase chain reaction (RT-PCR) assay of respiratory tract samples. Throat swabs were collected and maintained in the viral transport medium. The laboratory test assays for COVID-19 were conducted according to standards set by the World Health Organisation’s (WHO). Upper and lower respiratory tract specimens were collected in order to extract SARS-CoV-2 RNA. The RNA was obtained and further tested by means of RT-PCR using the same method that was described previously [20].

### 2.4 STATISTICAL ANALYSIS AND PLOTTING

Descriptive statistical analyses were performed to express categorical variables with numbers and proportions. These were then compared using a chi-square test. P values of less than or equal to 0.05 (two-sided) were considered statistically significant. R-script and GraphPad Prism version 7.04 was used to perform all of the statistical analyses and the figure plotting. Patients with at least one type of comorbidity were considered comorbid, and those with no comorbidity were considered non-comorbid patients.

## 3 RESULTS

### 3.1 EPIDEMIOLOGICAL FEATURES

Among the 168 COVID-19 patients admitted in the ICU with a confirmed outcome, 95 (56.5%) of the severely ill patients died in the ICU and the remaining 73 patients (43.5%) were transferred to the isolation ward following improvement (Table 1). Although 66.7% of the patients were over 50 years old, the highest proportion (28.6%) was between 51 and 60 years old. The proportion of male patients (79.8%) was more than female patients (20.2%). The COVID-19 individuals were into diverse professions and while the 10 (6.0%) of the patients had direct involvement in the healthcare system, most of the patients were from urban areas (65.5%). Persistence of a comorbidity was directly proportional to the state of being admitted in the ICU. As shown in Figure 1A, the proportion of deceased patients was relatively low in the group without comorbidities. Interestingly, the patients who were over 50 years old and had comorbidities comprise 66.3% of the total deaths, with the number of deaths being seven times the number of deaths in the group without comorbidities (Figure 1B). About 82.1% (138/168) of patients had at least one coexisting chronic illness, predominantly diabetes (52.4%), hypertension (41.1%) or heart disease (16.7%) (Table 1). The prevalence of diabetes, hypertension and heart disease in deceased patients was slightly higher (Figure 1C). Interestingly, patients with asthma survived well compared to other comorbidities. The most common symptoms experienced by patients were ARDS (133/168; 79.2%), fever (91/168; 54.2%) and coughing (43/168; 25.6%) (Figure 1D). The median length of hospital stay was five days, and the median length of ICU stay was four days (Figure 1E). The average duration of stay in the ICU was higher in surviving patients. In surviving patients, the median length of hospital stay was eleven days, and the median length of ICU stay was six days (Figure 1F). The leading causes of death were cardio respiratory failure (73/95; 76.8%), diabetes mellitus related complications (30/95; 31.6%), ARDS (20/95; 21.0%), pneumonia (20/95; 21.0%) and heart disease (7/95; 7.4%) (Figure 1G).

**Table 1:**
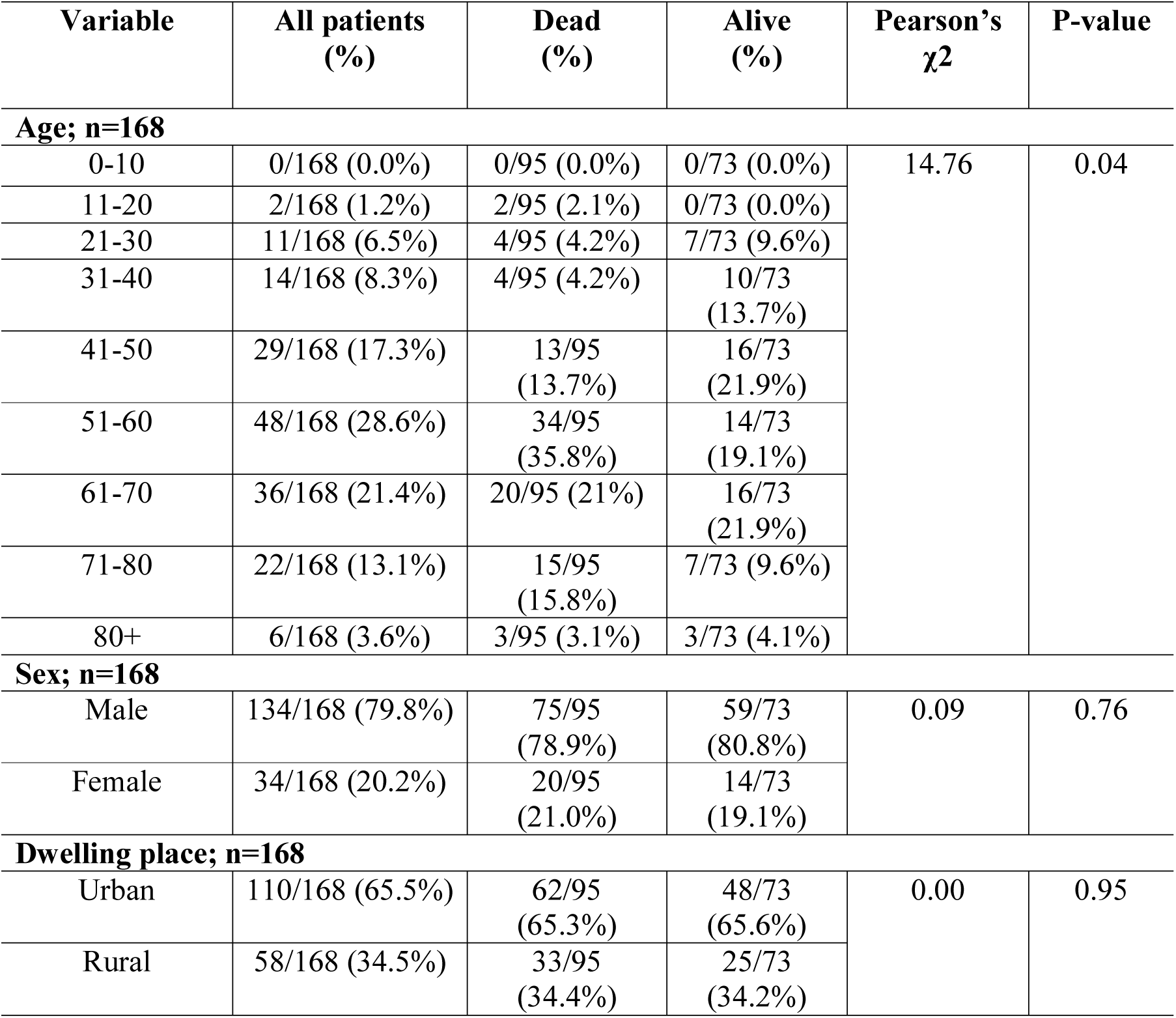
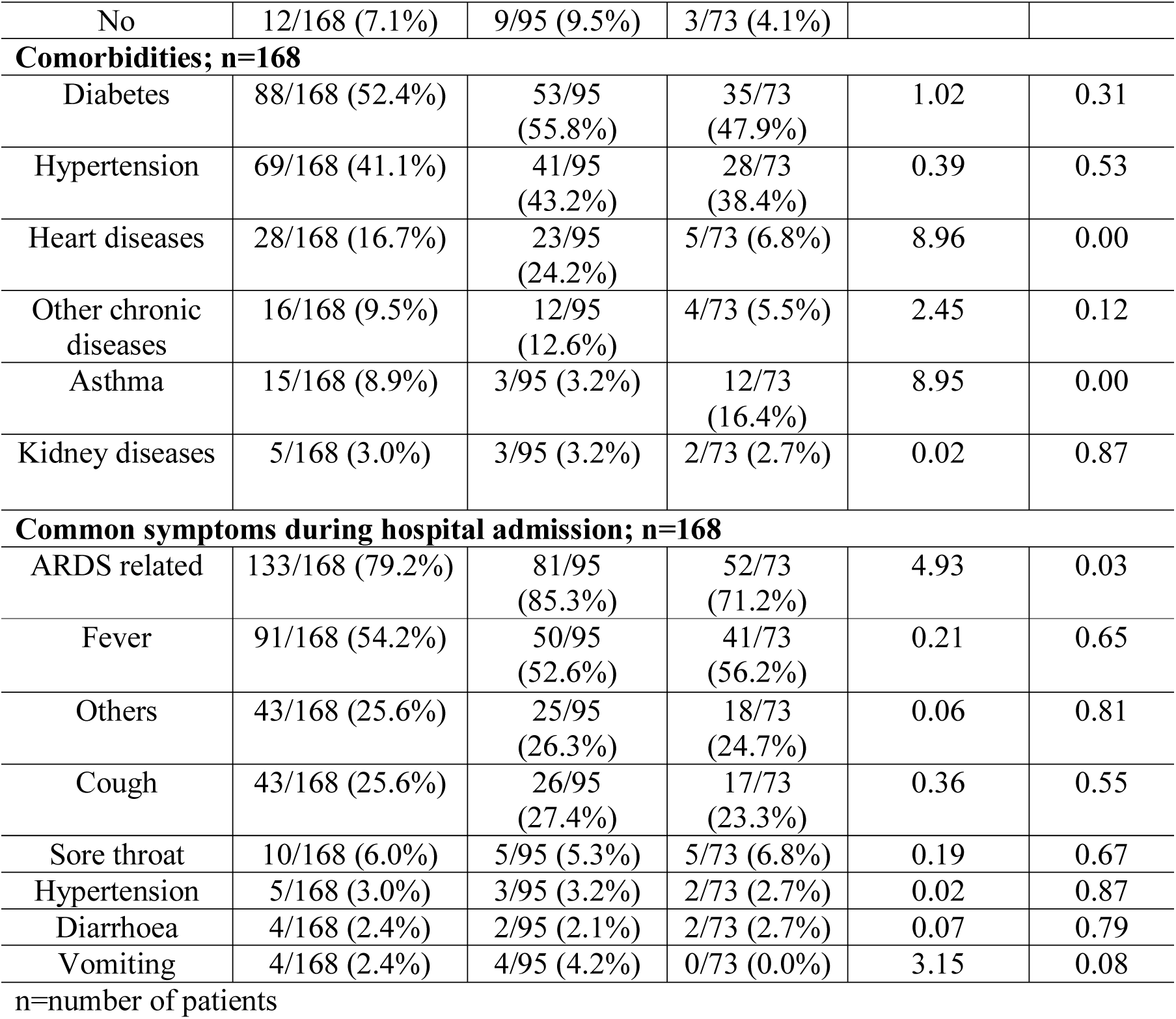
Demographic and baseline characteristics of COVID-19 ICU patients.

| Variable | All patients (%) | Dead (%) | Alive (%) | Pearson's $\chi^2$ | P-value |
| --- | --- | --- | --- | --- | --- |
| Age; n=168 |  |  |  |  |  |
| 0-10 | 0/168 (0.0%) | 0/95 (0.0%) | 0/73 (0.0%) | 14.76 | 0.04 |
| 11-20 | 2/168 (1.2%) | 2/95 (2.1%) | 0/73 (0.0%) |  |  |
| 21-30 | 11/168 (6.5%) | 4/95 (4.2%) | 7/73 (9.6%) |  |  |
| 31-40 | 14/168 (8.3%) | 4/95 (4.2%) | 10/73 (13.7%) |  |  |
| 41-50 | 29/168 (17.3%) | 13/95 (13.7%) | 16/73 (21.9%) |  |  |
| 51-60 | 48/168 (28.6%) | 34/95 (35.8%) | 14/73 (19.1%) |  |  |
| 61-70 | 36/168 (21.4%) | 20/95 (21%) | 16/73 (21.9%) |  |  |
| 71-80 | 22/168 (13.1%) | 15/95 (15.8%) | 7/73 (9.6%) |  |  |
| 80+ | 6/168 (3.6%) | 3/95 (3.1%) | 3/73 (4.1%) |  |  |
| Sex; n=168 |  |  |  |  |  |
| Male | 134/168 (79.8%) | 75/95 (78.9%) | 59/73 (80.8%) | 0.09 | 0.76 |
| Female | 34/168 (20.2%) | 20/95 (21.0%) | 14/73 (19.1%) |  |  |
| Dwelling place; n=168 |  |  |  |  |  |
| Urban | 110/168 (65.5%) | 62/95 (65.3%) | 48/73 (65.6%) | 0.00 | 0.95 |
| Rural | 58/168 (34.5%) | 33/95 (34.4%) | 25/73 (34.2%) |  |  |
| No | 12/168 (7.1%) | 9/95 (9.5%) | 3/73 (4.1%) |  |  |
| **Comorbidities; n=168** |  |  |  |  |  |
| Diabetes | 88/168 (52.4%) | 53/95 (55.8%) | 35/73 (47.9%) | 1.02 | 0.31 |
| Hypertension | 69/168 (41.1%) | 41/95 (43.2%) | 28/73 (38.4%) | 0.39 | 0.53 |
| Heart diseases | 28/168 (16.7%) | 23/95 (24.2%) | 5/73 (6.8%) | 8.96 | 0.00 |
| Other chronic diseases | 16/168 (9.5%) | 12/95 (12.6%) | 4/73 (5.5%) | 2.45 | 0.12 |
| Asthma | 15/168 (8.9%) | 3/95 (3.2%) | 12/73 (16.4%) | 8.95 | 0.00 |
| Kidney diseases | 5/168 (3.0%) | 3/95 (3.2%) | 2/73 (2.7%) | 0.02 | 0.87 |
| **Common symptoms during hospital admission; n=168** |  |  |  |  |  |
| ARDS related | 133/168 (79.2%) | 81/95 (85.3%) | 52/73 (71.2%) | 4.93 | 0.03 |
| Fever | 91/168 (54.2%) | 50/95 (52.6%) | 41/73 (56.2%) | 0.21 | 0.65 |
| Others | 43/168 (25.6%) | 25/95 (26.3%) | 18/73 (24.7%) | 0.06 | 0.81 |
| Cough | 43/168 (25.6%) | 26/95 (27.4%) | 17/73 (23.3%) | 0.36 | 0.55 |
| Sore throat | 10/168 (6.0%) | 5/95 (5.3%) | 5/73 (6.8%) | 0.19 | 0.67 |
| Hypertension | 5/168 (3.0%) | 3/95 (3.2%) | 2/73 (2.7%) | 0.02 | 0.87 |
| Diarrhoea | 4/168 (2.4%) | 2/95 (2.1%) | 2/73 (2.7%) | 0.07 | 0.79 |
| Vomiting | 4/168 (2.4%) | 4/95 (4.2%) | 0/73 (0.0%) | 3.15 | 0.08 |

n=number of patients

**Table 2:**
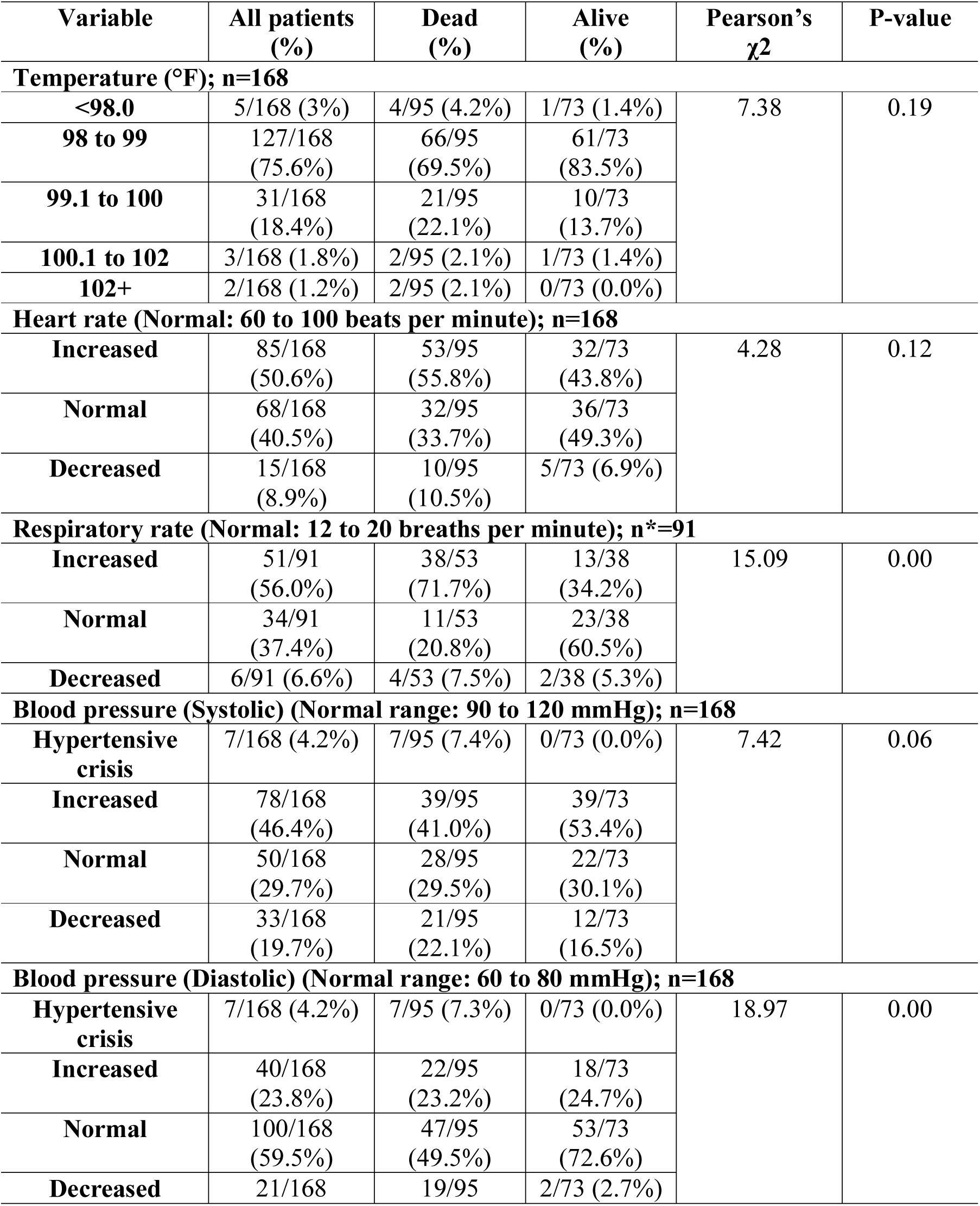
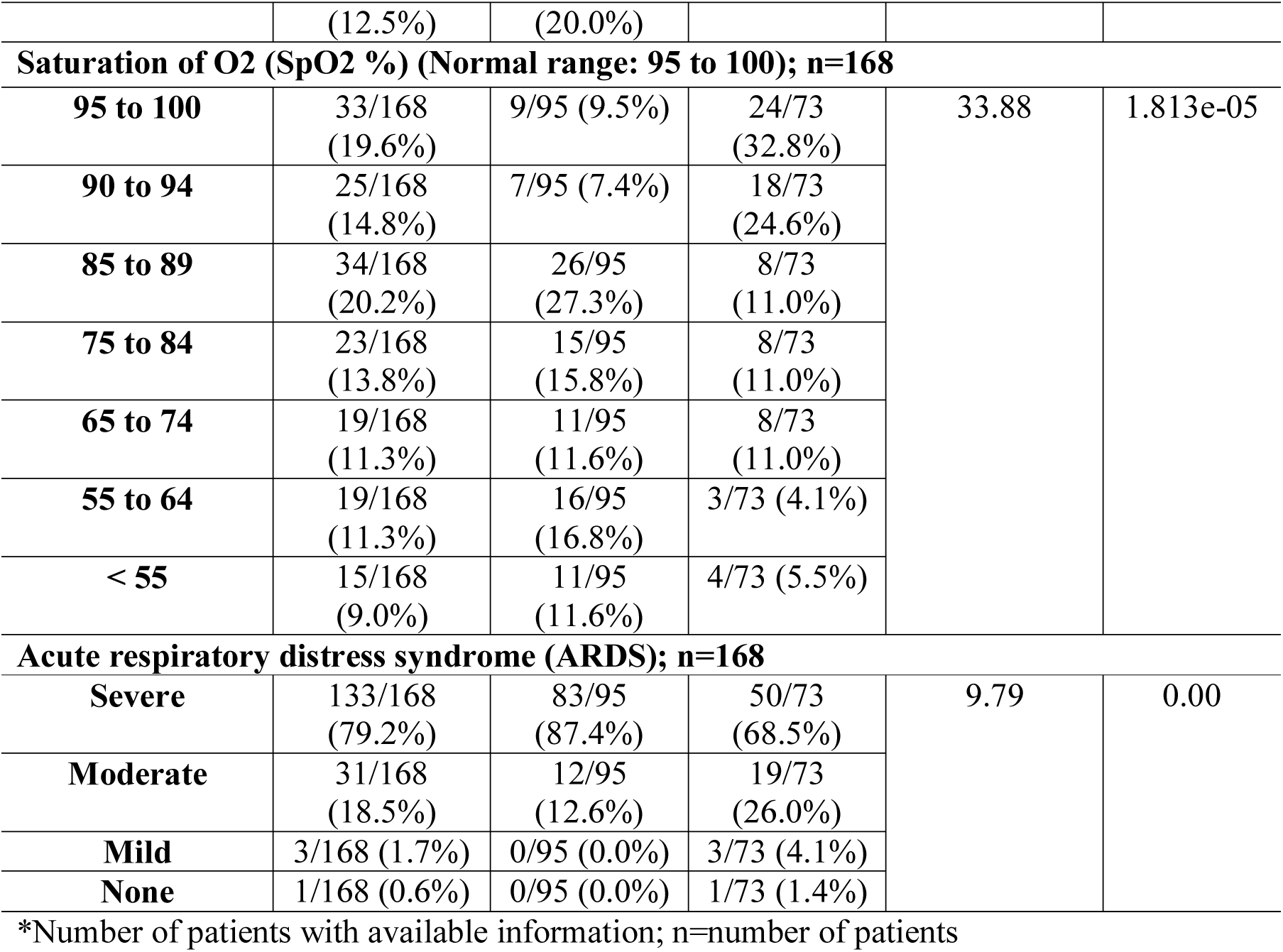
Vital signs at ICU admission of COVID-19 patients.

**Figure 1:**
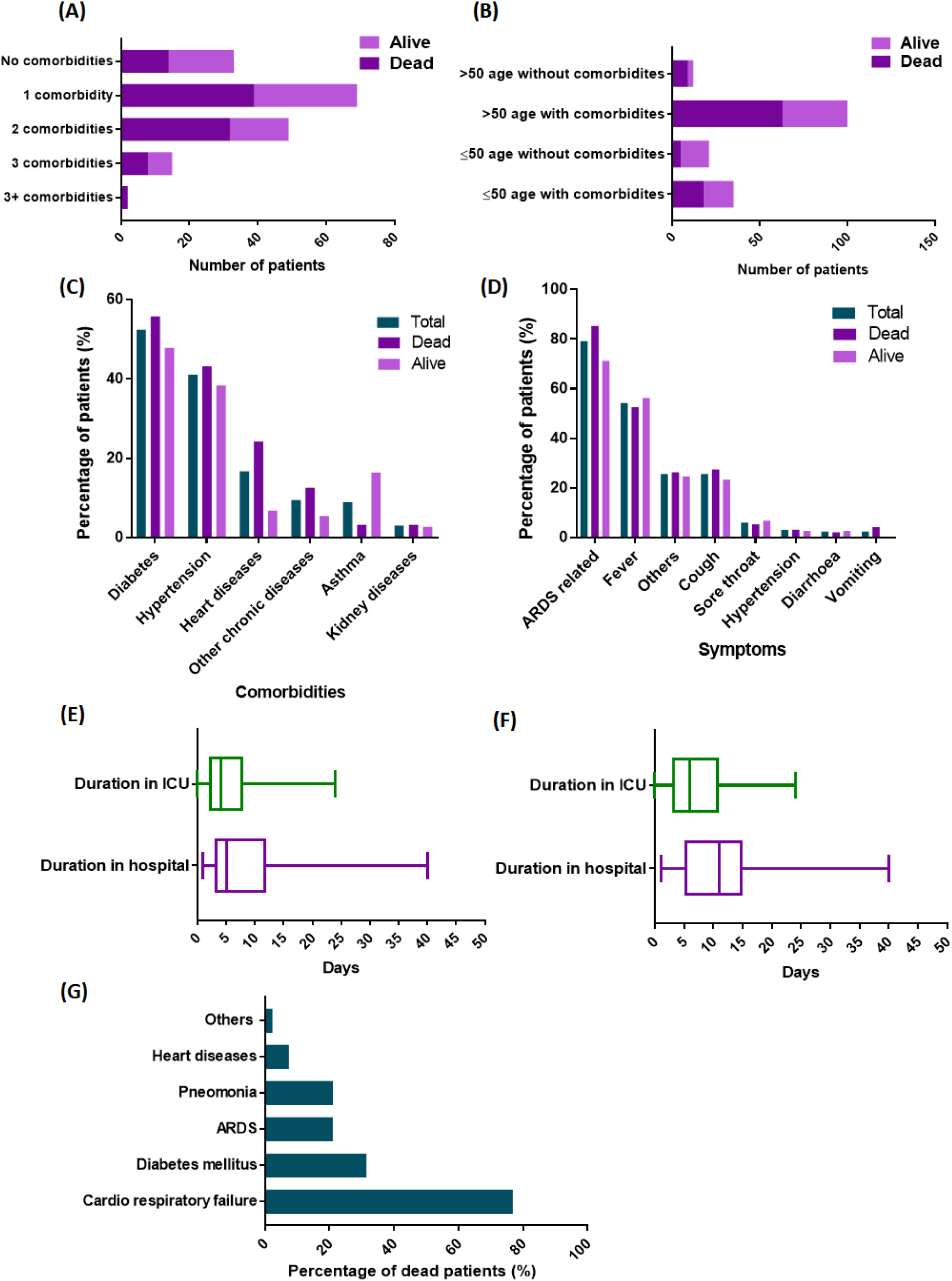
The clinical features of Bangladeshi patients infected with COVID-19 admitted to the ICU. (A) Frequency of number of comorbidities in patients admitted to the ICU; (B) Relationship between age and comorbidities and its frequency in patients; (C) Percentage of the occurrence of different comorbidities in total, dead and alive patients; (D) Percentage of the occurrence of different symptoms in total, dead and alive patients; (E) Boxplot of the number of days in the ICU and hospital for patients admitted to the ICU, with the boxes spanning the 25 to 75 percentiles and the horizontal lines in the boxes representing the medians; (F) Boxplot of the number of days in the ICU and hospital for survived patients admitted to the ICU, with the boxes spanning the 25 to 75 percentiles and the horizontal lines in the boxes representing the medians;(G) Distribution of the reasons for death. ARDS: acute respiratory distress syndrome; ICU: intensive care unit

### 3.2 VITAL SIGNS AND PHYSICAL EXAMINATION

The body temperatures for all individuals in the study sample were measured, and this ranged from 98°F to 102+°F. The vital signs at admission to the ICU were moderate fever ≥ 99°F for 40 patients (71.1%), heart rate ≥ 100 beats per minute for 85 patients (51%) and a respiratory rate of ≥ 25 breaths per minute in 56% of the recorded patients. The patients who had a moderate or high fever (≥ 99°F) tended to have a higher mortality rate than those with a mild or no fever (Figure 2A). Oxygen saturation was low (< 95% SpO2) in 135 patients (80.4%) and the mortality rate of these patients was relatively high (Figure 2B). The death rate of patients who had an abnormal heart rate and respiratory rate was higher (Figure 2C). Shock occurred in 10 patients (5.9%), including cardiogenic shock in five patients (2.9%) and septic shock in four patients (2.4%) (Figure 2D). Of all 168 ICU patients, seven (4.2%) were classified as having a hypertensive crisis. Unfortunately, none of these patients survived. ARDS occurred in 167 (99.4%) patients, with 31 patients (18.5%) having moderate ARDS, three patients (1.8%) having mild ARDS and 133 patients (79%) experiencing severe ARDS. Eighty three out of the 133 severe ARDS patients (62.4%) died (Figure 2E).

**Figure 2:**
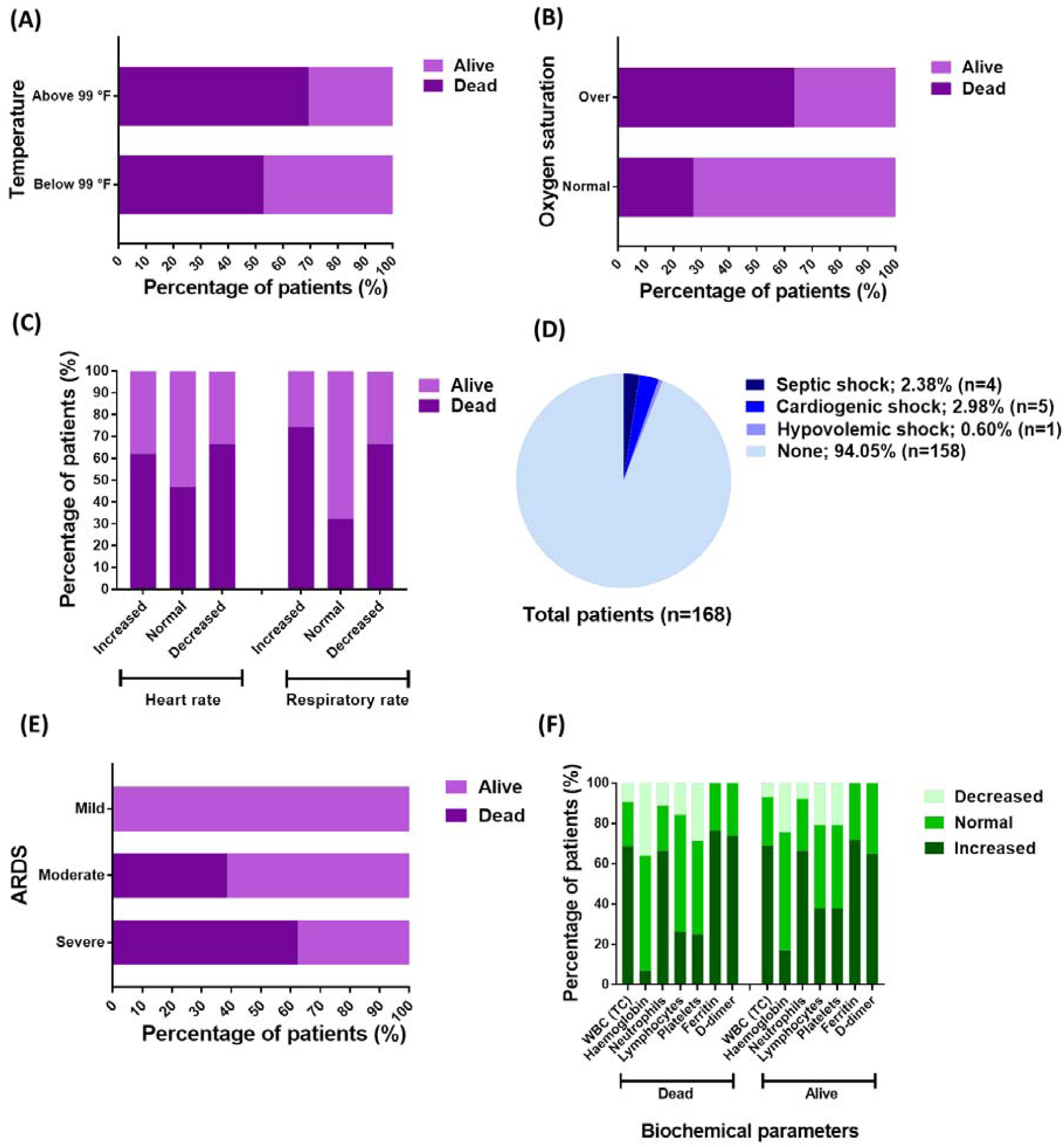
Vital signs, risk factors and laboratory findings. (A) Distribution of the clinical outcomes (dead or alive) of the patients in the no or low fever (≤ 99°F) group compared to those in the moderate or high fever (> 99°F) group; (B) Distribution of the clinical outcomes (dead or alive) of the patients in the normal oxygen saturation (≥ 95% SpO2) group compared to those in the low oxygen saturation (< 95% SpO2) group; (C) Distribution of dead and alive patients according to their heart rate and respiratory rate, namely whether it was increased, normal or decreased, with the normal reference values being a heart rate of 60 to 100 beats per minute and a respiratory rate of 12 to 20 breaths per minute; (D) Pie chart of the occurrence of septic shock, cardiogenic shock and hypovolemic shock; (E) Distribution of the clinical outcomes (dead or alive) of patients in the mild, moderate and severe ARDS groups; (F) Biochemical parameters of COVID-19 patients admitted to the ICU, with the normal reference values being: normal range of WBC of 4–10×10^9^ per L, normal range of haemoglobin of 130–175g per L, normal range of neutrophils of 1.8–6.3×10^9^ per L, normal range of lymphocytes of 1.1–3.2×10^9^ per L, normal range of platelets of 125–350×10^9^ per L, D-dimer < 0.5 µg/mL, Ferritin < 500 μg/L. ARDS: acute respiratory distress syndrome; F: Fahrenheit; ICU: intensive care unit; L: litre; TC: total count; WBC: white blood cell.

### 3.3 LABORATORY FINDINGS

The laboratory findings of the patients upon admission to the ICU are shown in Figure 2 and Table 3. Statistical analysis was only conducted on the patients whose laboratory results were available. Elevated levels of White Blood Cell (WBC) and Neutrophils were identified in 68.9% (42/61), and 66.7% (44/64) patients, respectively. For 71 patients who underwent tests on D-dimer, an excessive level was identified from 49 (69.0%) patients, with the level higher than 5 mg/L in 15 (21.1%) patients. Out of the 69 patients who had tests of Ferritin, elevated levels of Ferritin were identified in 53 (76.8%) patients (Table 3). The biochemical parameters of the survived and non-survived patients were also compared, and it was found that they were essentially identical (Figure 2F).

**Table 3:** Laboratory findings of patients with COVID-19 in ICUs.

| Variable | All patients (%) | Dead (%) | Alive (%) | Pearson's $\chi^2$ | P-value |
| --- | --- | --- | --- | --- | --- |
| **White blood cell count ($\times 10^9$ per L; normal range 4–10); n\*=61; Dead\*=32 ; Alive\*=29** |  |  |  |  |  |
| Increased | 42/61 (68.9%) | 22/32 (68.8%) | 20/29 (69.0%) | 0.15 | 0.93 |
| Normal | 14/61 (23.0%) | 7/30 (21.9%) | 7/29 (24.1%) |  |  |
| Decreased | 5/61 (8.2%) | 3/30 (9.4%) | 2/29 (6.9%) |  |  |
| **Haemoglobin (g/L; normal range 130–175); n\*=57; Dead\*=28; Alive\*=29** |  |  |  |  |  |
| Increased | 7/57 (12.3%) | 2/28 (7.1%) | 5/29 (17.2%) | 1.82 | 0.40 |
| Normal | 33/57 (57.9%) | 16/28 (57.1%) | 17/29 (58.6%) |  |  |
| Decreased | 17/57 (29.8%) | 10/28 (35.7%) | 7/29 (24.1%) |  |  |
| **Neutrophils ($\times 10^9$ per L; normal range 1.8–6.3); n\*=66; Dead\*=27; Alive\*=39** |  |  |  |  |  |
| Increased | 44/64 (66.7%) | 18/27 (66.7%) | 26/39 (66.7%) | 0.28 | 0.87 |
| Normal | 16/64 (24.2%) | 6/27 (22.2%) | 10/39 (25.6%) |  |  |
| Decreased | 6/64 (9.1%) | 3/27 (11.1%) | 3/39 (7.7%) |  |  |
| **Lymphocytes ($\times 10^9$ per L; normal range 1.1–3.2); n\*=48; Dead\*=19; Alive\*=29** |  |  |  |  |  |
| Increased | 16/48 (33.3%) | 5/19 (26.3%) | 11/29 (37.9%) | 1.27 | 0.53 |
| Normal | 23/48 (47.9%) | 11/19 (57.9%) | 12/29 (41.4%) |  |  |
| Decreased | 9/48 (18.8%) | 3/19 (15.8%) | 6/29 (20.7%) |  |  |
| **Platelets ($\times 10^9$ per L; normal range 125–350); n\*=57; Dead\*=28; Alive\*=29** |  |  |  |  |  |
| Increased | 18/57 (31.6%) | 7/28 (25.0%) | 11/29 (37.9%) | 1.20 | 0.55 |
| Normal | 25/57 (43.9%) | 13/28 (46.4%) | 12/29 (41.4%) |  |  |
| Decreased | 14/57 (24.6%) | 8/28 (28.6%) | 6/29 (20.7%) |  |  |
| **D-dimer (mg/L; normal range < 0.5); n\*=71; Dead\*=31; Alive\*=40** |  |  |  |  |  |
| Normal | 22/71 (31.0%) | 8/23 (25.8%) | 14/48 (35.0%) | 2.4 | 0.48 |
| > 0.5 to ≤ 5 | 34/71 (47.9%) | 16/23 (51.6%) | 18/48 (45.0%) |  |  |
| > 5 to ≤ 10 | 11/71 (15.5%) | 4/23 (12.9%) | 7/48 (17.5%) |  |  |
| > 10 | 4/71 (5.6%) | 3/23 (9.7%) | 1/48 (2.5%) |  |  |
| **Ferritin concentration (µg/L; normal range < 500); n\*=69; Dead\*=30; Alive\*=39** |  |  |  |  |  |
| Normal | 16/69 (23.2%) | 7/30 (23.3%) | 9/39 (28.1%) | 1.65 | 0.80 |
| ≥ 500 to < 1000 | 25/69 (36.2%) | 13/30 (43.3%) | 12/39 (30.8%) |  |  |
| **≥ 1000 to < 1500** | 13/69 (18.8%) | 5/30 (16.7%) | 8/39 (20.5%) |  |  |
| **≥ 1500 to < 2000** | 11/69 (15.9%) | 4/30 (13.3%) | 7/39 (17.9%) |  |  |
| **≥ 2000** | 4/69 (5.8%) | 1/30 (3.3%) | 3/39 (7.7%) |  |  |

\*Number of patients with available information; ICU, Intensive care unit; L, Litre; n=number of patients

### 3.4 MANAGEMENT AND MEDICATIONS

Oxygen therapy was administered in accordance with the patients’ oxygen saturation. Over 90% of the patients who were admitted to the ICU (158/168; 93.6%) required oxygen during the course of the disease. Of the 158 patients with available information, 64.8% (103/159) received oxygen support via a mask, and 25.2% (40/159) received oxygen support via a high flow nasal cannula. Prone positioning was implemented to enhance oxygenation and improve lung recruitability in some patients with severe ARDS (151/168; 89.9%) (Table 4). Convalescent plasma (CP) was transfused into eight patients. However, only three of these eight patients survived after the convalescent plasma transfusion. As for the medications administered, 94 patients (56.0%) received antiviral agents, 164 patients (97.6%) received antimicrobial agents, 126 patients (75.0%) received an anti-allergic drug, 149 patients (88.7%) received anti-inflammatory drugs and 145 patients (86.3%) received vitamin and mineral supplements. Favipiravir (71/168; 42.3%) and Remdesivir (31/168; 18.4%) were the most commonly used antiviral drugs among the ICU patients. However, the proportion of surviving patients was greater in the Remdesivir cohort than the Favipiravir cohort (Figure 3A). Additionally, Methylprednisolone (97/168; 57.7%) and Dexamethasone (40/168; 23.8%) were the two most used glucocorticoids (Figure 3A). Meropenem (126/168; 76.2%) was the most commonly used antibiotic, followed by Ceftriaxone (50/168; 29.8%), Azithromycin (33/168; 19.4%) and Moxifloxacin (33/168; 19.4%) (Figure 3A). As shown in Figure 3B, the proportion of survived patients was slightly higher with the use of Meropenem, as well as Remdesivir and Dexamethasone, than with the use of Favipiravir or Methylprednisolone. Six patients were treated with Remdesivir and Dexamethasone and only one of them died. The vitamin C, vitamin D and zinc supplements that were commonly used did not show any improved clinical outcomes (Figure 3C).

**Table 4:**
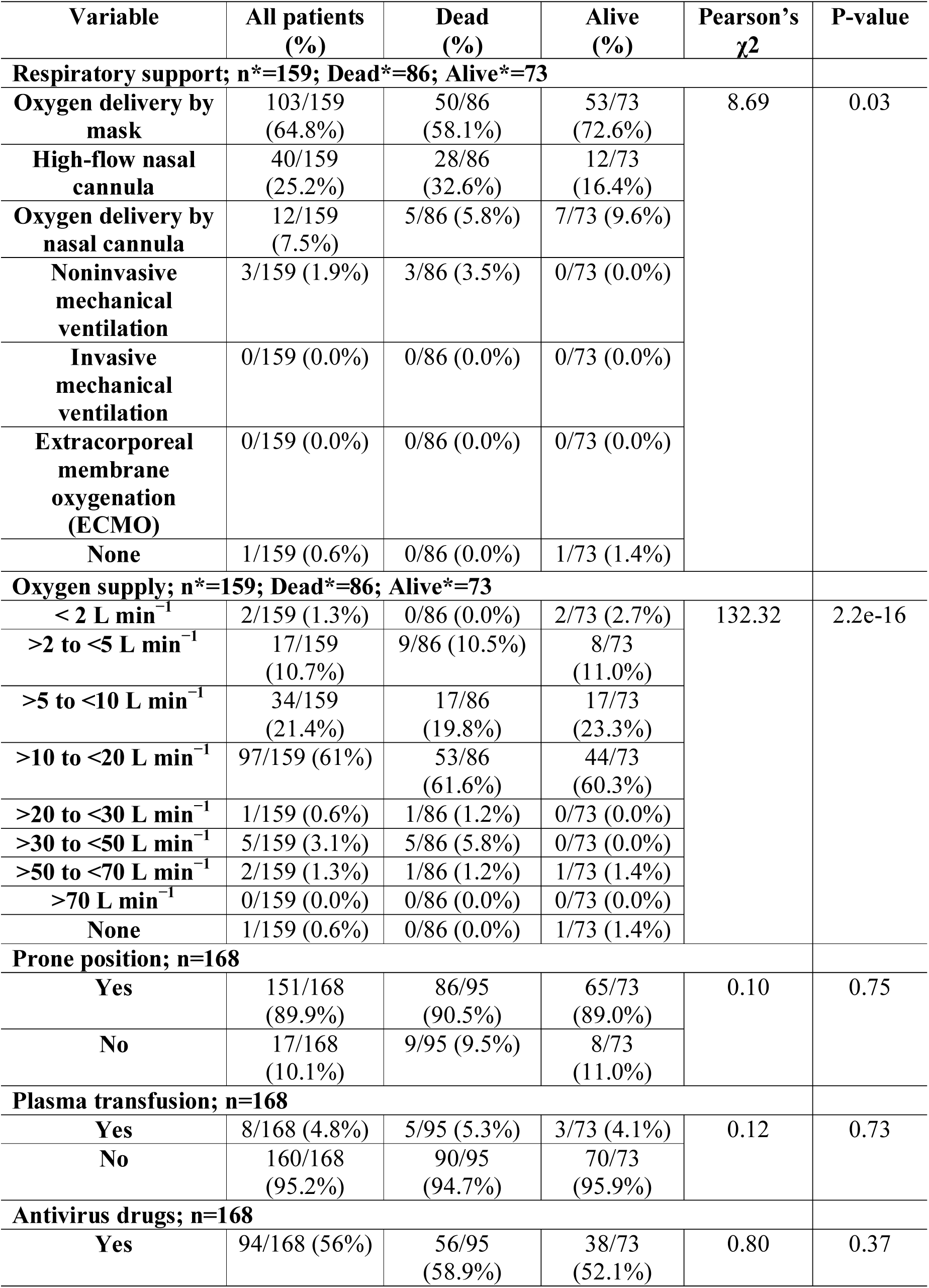
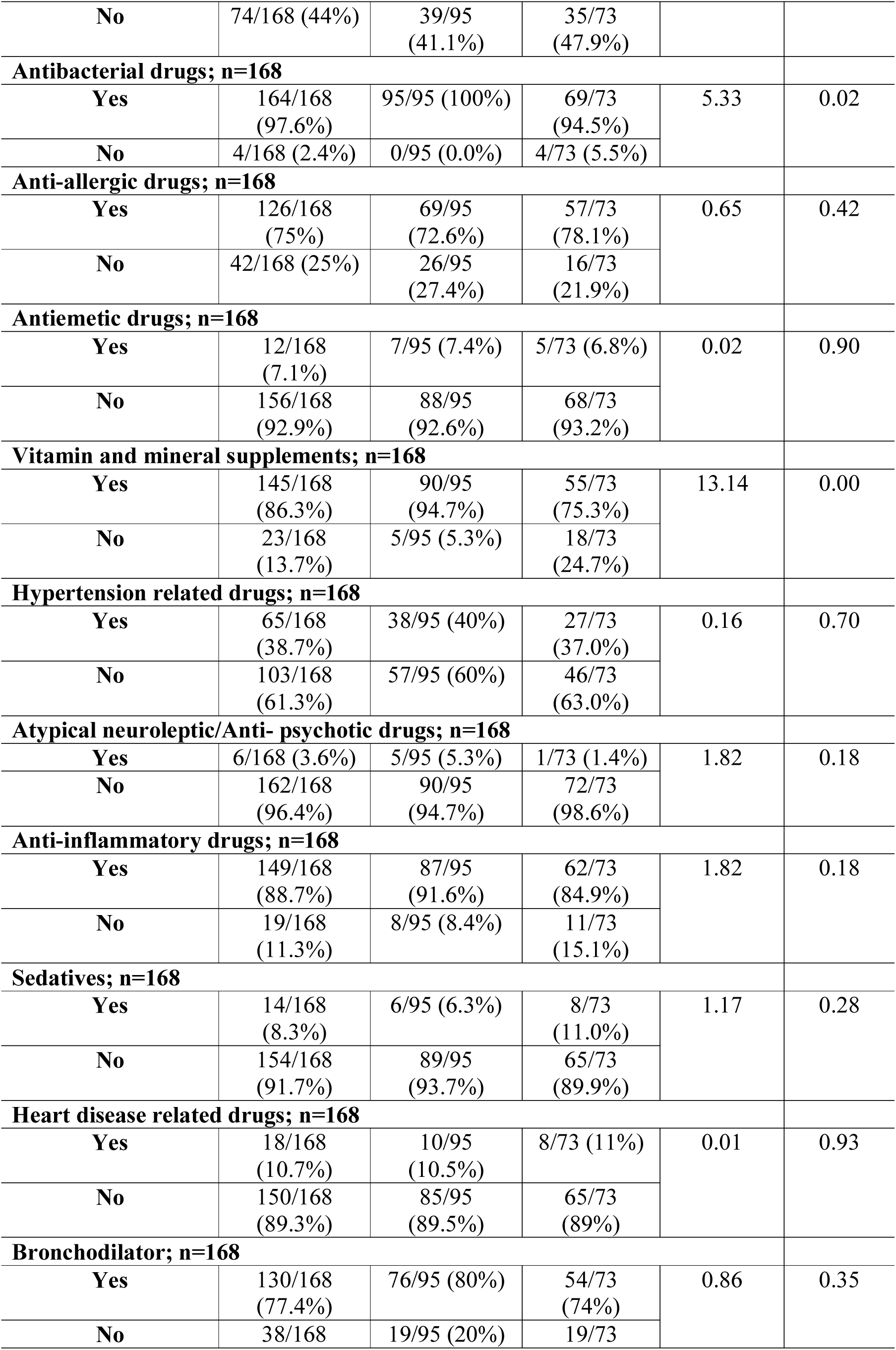
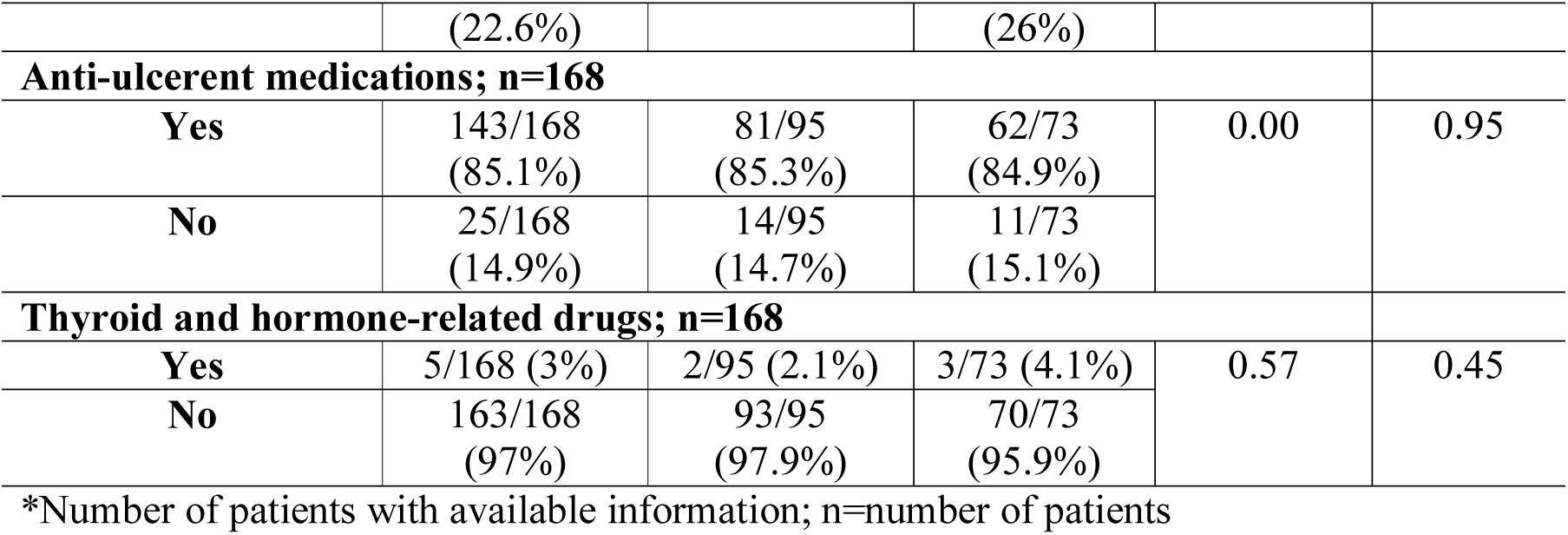
Managements of patients with COVID-19 in ICUs.

**Figure 3:**
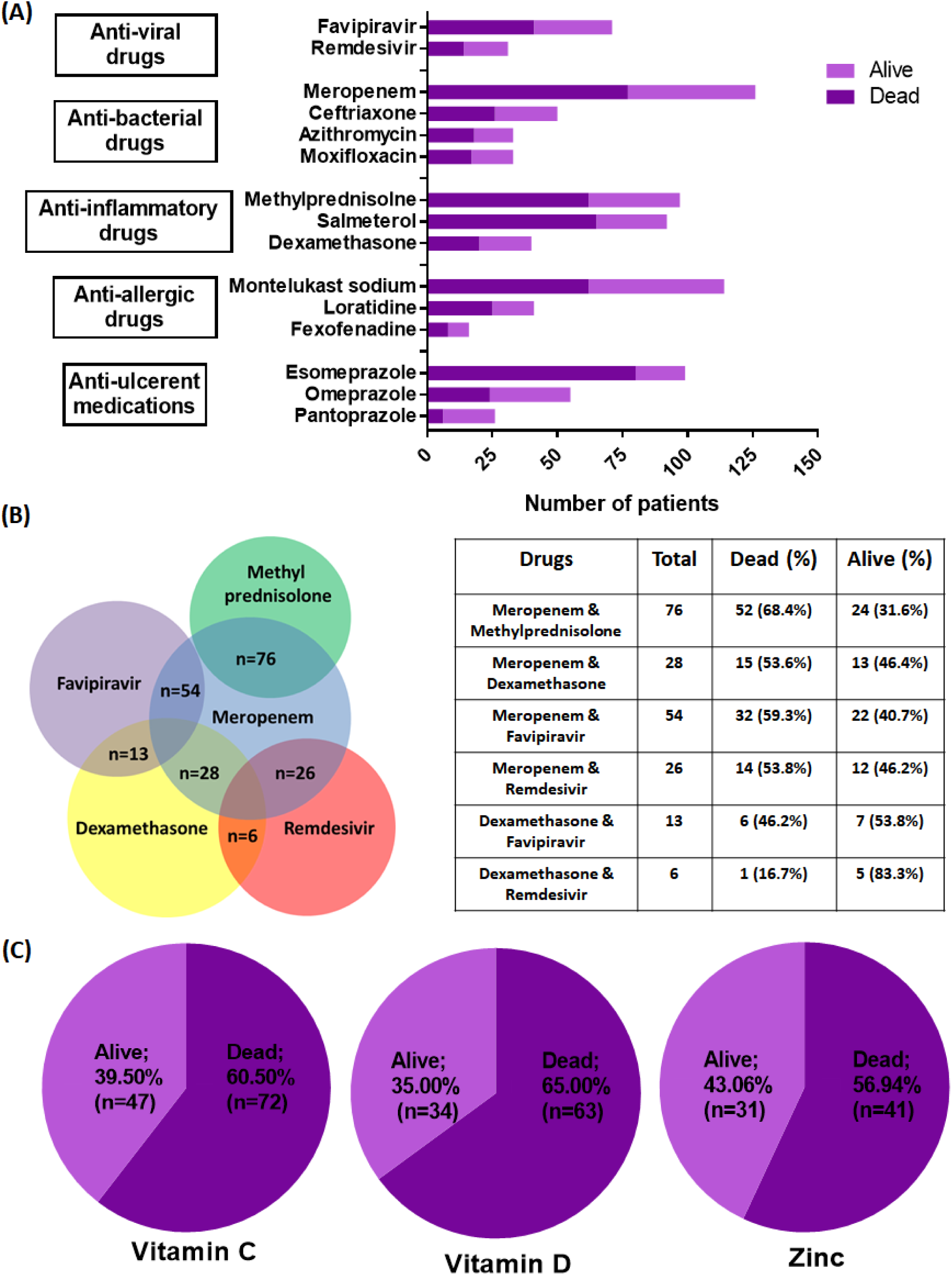
The medicines, vitamins and electrolytes commonly administered to COVID-19 patients in the ICU. (A) Different generics of the drugs administer to COVID-19 patients in the ICU; (B) Venn diagram of drug combinations for the commonly used antibiotic, Meropenem with two antiviral drugs and two glucocorticoids, with the antiviral drugs being Favipiravir and Remdesivir and the glucocorticoids being Methylprednisolone and Dexamethasone and the proportions of dead and alive patients in each overlapped drug group being shown as a percentage; (C) Pie chart illustrating the numerical proportion of dead and alive patients who took Vitamin C, Vitamin D and Zinc supplements.

## 4 DISCUSSION

On 22^nd^ September 2020, COVID-19 cases in Bangladesh totalled to 352,178, with 260,790 recovered cases and 5007 deaths. The information on the clinical characteristics of COVID-19 individuals having an advanced and deleterious prognosis of COVID-19 is still scarce although the positive cases are nowhere near decreasing. The median age of the critical COVID-19 patients of the sample in this study (56 years) is lower than that of Italy, the United States of America (USA), Greece and China [21-24]. However, the gender propensity of this study’s patients (mostly men) is consistent with that of COVID-19 patients in ICUs in Italy, USA and China [21-23]. The management of patients with several comorbidities is challenging due to their frailty and increased risk of mortality, which is amplified when these comorbid individuals are diagnosed with COVID-19. The current study has found that older (≥ 50) Bangladeshi male patients with previous comorbidities, such as diabetes, hypertension and heart diseases, are profoundly susceptible to COVID-19, which is comparative to the pattern that has been revealed in China, Italy and New York [8, 21, 23, 25]. In Bangladesh, most people diagnosed with diabetes are from urban areas, and the prevalence of diabetes is highest among those aged from 55 to 59 years [26]. The presence of comorbidity might explain COVID-19’s severity in Bangladeshi patients aged 51 to 60 years.

Another diverting finding from this study was that patients with asthma survived well compared to other comorbidities. As with other viruses, SARS-CoV-2 triggers asthma exacerbations, which is why asthma is listed as a risk factor for COVID-19 related morbidity. However, this study’s finding is consistent with that of Leonardo Antonicelli et al. (2020), who found that asthma seems to play a minimal role in clinical severity [27]. ARDS (79.2%) was found to be the most prominent symptom within the study sample upon admission to the ICU, and this was also reflected in patients described in reports from China, USA and Europe [8, 21, 22]. Other noteworthy symptoms are fever (8.40%) and coughing (7.70%), and the results obtained by this study align with the trends concerning high prevalence seen in other countries [21-23]. Intestinal signs and symptoms, such as diarrhoea, were rarely developed by the patients in this study. The vast majority of this study’s patients needed oxygen support upon admission to the ICU due to severe to moderate ARDS as indicated by their extremely low oxygen saturations. This study finds that there is a high mortality rate among patients with a moderate to high fever and a low oxygen saturation.

Therapeutic plasma exchange has been recommended as a treatment measure for patients with severe COVID-19; however, this study found that therapeutic plasma exchange had no significant impact on the improvement of critically ill patients. According to a recent study, therapeutic plasma exchange can be effective in critically ill patients if it can be applied within the first week of symptom onset [28]. Unfortunately, most of the patients in the current study were admitted to the ICU in a critical condition due to the lack of available ICU beds. Therefore, it may have been too late for convalescent plasma therapy to have an effective impact.

To the extent of the author’s knowledge, so far, this study is the only study on the medicine administered to critically ill COVID-19 patients in Bangladesh. Currently, there is no recommended treatment for COVID-19 infection in careful supportive care [29]. In this study, 97.6% of patients received antibacterial agents, 56% received antiviral therapy and 88.7% received anti-inflammatory drugs. Even though the antiviral drug Favipiravir was the mostly used antiviral drug, the survival rate was higher among the patients who had been given Remdesivir. Favipiravir concentrations become lower in critically ill patients than in healthy subjects, which might be one reason why Favipiravir is less effective [30]. Several countries, such as Japan, Taiwan and USA, and the European Union (EU) suggest the conditional use of Remdesivir to treat critical patients [31, 32]. Therefore, Remdesivir can be a better choice over Favipiravir in providing aid to COVID-19 individuals.

A recent report suggests that glucocorticoids may also minimize severe clinical outcomes in critical COVID-19 patients with ARDS [33]. The current study finds that Dexamethasone has comparatively better clinical outcomes than Methylprednisolone. According to a large clinical trial conducted in the United Kingdom (UK), Dexamethasone reduced deaths by about one-third in critical COVID-19 patients who were on ventilator support [34]. In this study, only one out of six patients who were treated with both Remdesivir and Dexamethasone died. However, further studies with larger sample sizes are required to evaluate the effectiveness of the combined use of Remdesivir and Dexamethasone.

Although the findings of this study were significant, limitations were also in order. Firstly, laboratory data collection to conduct a broad and extensive study was inevitably challenging as the laboratory results were not systematically collected. Secondly, the evaluated data was extracted retrospectively from patients’ medical files and not all laboratory tests were conducted on all patients. Thirdly, because of the study’s objective to identify the critical care needs of patients with the greatest severity of illness, the sample size is small. Therefore, more thorough assessment of comorbidities in larger samples of critical Bangladeshi patients with COVID-19 and future studies are required. Despite these limitations, this study represented the largest cohort of critically ill COVID-19 patients from Bangladesh reported to date.

## 5 CONCLUSION

To summarize, parallel to the data obtained from studies conducted in other countries, there is an elevated prevalence of comorbidities, such as diabetes, hypertension and heart diseases, in a profuse number COVID-19 patients with critical expositions who are hospitalised in Bangladesh. Since this cohort is more vulnerable in terms of COVID-19 related morbidity and mortality, besides implementing an effective policy for the prevention and control of the disease in general, the authorities should pay more attention to these atypical patients. In conclusion, the findings reported here provide important context for effective strategies for the provision of comprehensive health care to critically ill COVID-19 patients. However, future studies with larger sample sizes are needed in order to assess the risk factors and associated clinical outcomes in a broader sense.

## Data Availability

Not applicable

## 6 SUPPLEMENTARY FIGURES

**Supplementary Figure 1:**
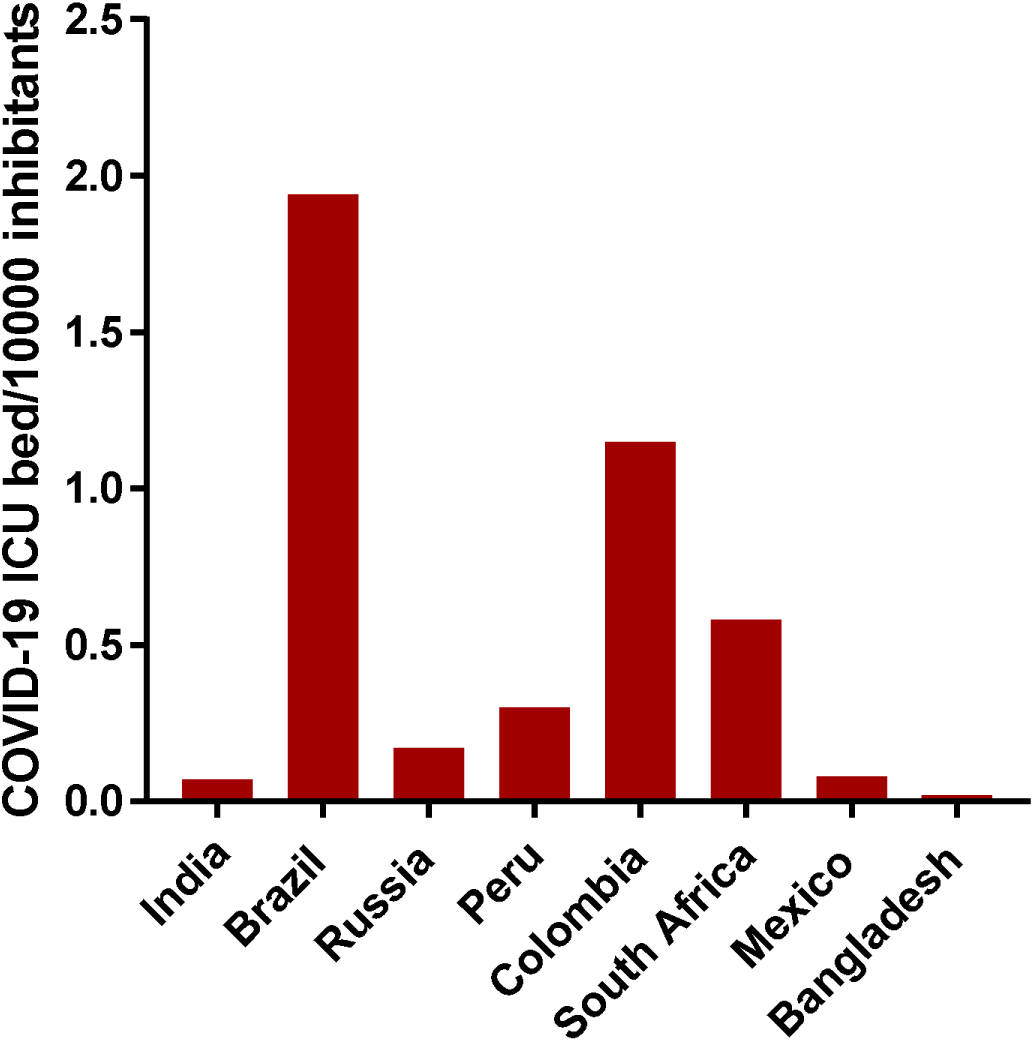
The number of COVID-19 ICU beds available for every 10,000 inhabitants of Bangladesh and other countries worse affected by COVID-19 [35-41].

## Abbreviations

ARDS: acute respiratory distress syndrome
COVID-19: Coronavirus disease 2019
ECMO: extracorporeal membrane oxygenation
ICU: intensive care unit
RT-PCR: Real-time reverse transcription polymerase chain reaction
WHO: World Health Organization

## Author contributions

AS, MMA, HMM, AP and TUQ: Study design. SN, ASA, AUC, HK, TR and PD Literature review, Data collection. MUSS, HMM and AS Data analysis, Visualization. SN, HMM and AS Manuscript writing, Editing. All authors reviewed and approved the final manuscript paper; approved the final version and agreed to be accountable for the work.

## Code availability

The source code and pipeline to reproduce our analyses can be accessed at https://github.com/sharifshohan/Cross_Sectional_Study_Bangladesh.

## Acknowledgements

The authors would like to acknowledge all patients and fellow healthcare workers for providing excellent patient care at considerable personal risk.

## Notes

### Competing Interest Statement

The authors have declared no competing interest.

### Author Declarations

Ethical committee of General hospital Chittagong, Chattogram, Bangladesh

